# Gut microbiome signatures associated with self-reported allergic symptoms among Finnish adults

**DOI:** 10.64898/2026.07.03.26357002

**Authors:** Himmi Lindgren, Ville Vartiainen, Geraldson Muluh, Nitin Bayal, Katariina Pärnänen, Guillaume Meric, Pekka Jousilahti, Matti Ruuskanen, Rob Knight, Teemu Niiranen, Aki S. Havulinna, Veikko Salomaa, Pande Putu Erawijantari, Leo Lahti

## Abstract

**Background:** Growing evidence suggests that the gut microbiome influences nasal and ocular allergic inflammation through gut-mucosal immune interactions. Yet, its association with Allergic rhinitis (AR) and allergic eye symptoms (AES) remains incompletely understood in large population-based cohorts.

**Objective:** To examine associations between the gut microbiome and self-reported AR and AES in Finnish adults.

**Methods:** Shallow metagenomic sequencing was performed on stool samples from a population-based cohort (FINRISK02; n = 7,231). Microbial taxonomic and functional profiles were compared between individuals with AR (n = 1,950), AES (n = 1,554), combined allergies (AR and/or AES; n = 2,305), and controls without reported symptoms (n = 3,175).

**Results:** Allergic groups exhibited lower microbial richness and phylogenetic diversity than controls. Shared microbial and functional signatures were observed across AR and AES, consistent with their high co-occurrence (N = 1,199). Compared with controls, allergic groups showed enrichment of 17 bacterial species, predominantly from the *Clostridia* class, including taxa previously associated with asthma, chronic obstructive pulmonary disease, and atopic dermatitis. Allergic individuals also exhibited enrichment of pathways related to mucosal carbohydrate processing, shikimate metabolism, histidine turnover, and broader amino acid metabolism. Concurrent enrichment of histidine biosynthesis and degradation suggested altered microbial histidine metabolism.

**Conclusions:** Adult allergic symptoms are associated with gut microbiome taxonomic and functional alterations linked to mucosal barrier function and immune-related metabolism, supporting a shared gut-mucosal immune axis across allergic phenotypes.

**Clinical Implication:** Shared microbial signatures across AR and AES support the concept of a common gut-mucosal immune axis underlying allergic manifestations in adults.

**Capsule Summary:** The large population-based study revealed that allergic symptoms in adults are associated with lower richness and phylogenetic diversity of gut microbiome together with taxonomic and functional alterations

## Introduction

Allergic rhinitis (AR) and allergic eye symptoms (AES) are common allergic conditions with increasing prevalence among adults^1,2^. AR often coexists with AES due to shared underlying pathophysiological mechanisms, as both conditions are part of a broader IgE-mediated allergic response driven by T-helper type 2 (TH2) immunity and influenced by environmental and genetic factors^3^. Consistent with this mechanism, large European cohort studies^4–6^ have reported that more than 70% of individuals with AR also experience AES.

Growing evidence suggests that the gut microbiome contributes to immune regulation and the development of allergic disease^7–9^. Reduced exposure to environmental microbiota has been proposed to impair immune tolerance and increase susceptibility to allergic disorders, as described by the hygiene and biodiversity hypotheses^10–12^. In support of this concept, studies from Karelia demonstrated lower allergy prevalence in the Russian population compared with the Finnish population, despite the study populations being located only ∼200 km apart and sharing genetic ancestry and climate, alongside richer microbial exposures and more balanced immune regulatory networks^11,12^. Microbial diversity has also been proposed to play a crucial role in shaping immune tolerance and preventing overactive immune responses seen in conditions like AR and other allergic conditions^13,14^. Alterations in gut microbiota have been reported to be associated with several allergic diseases, such as atopic dermatitis, seasonal allergies, and food allergies^13–16^. While there have been limited studies on the association between gut microbiome and AR^17^, recent studies have begun to explore the association between gut microbiome and AES^18^. Thus, more research is needed to understand the links between the gut microbiome and, in particular, the long-term impact on chronic allergy symptoms.

The FINRISK 2002 microbiome study^20,21^ provides a unique opportunity to investigate associations between gut microbiome and allergy in a large cohort of Finnish adults. We compared taxonomic diversity, composition, and predicted microbial functions between individuals with allergic symptoms (AR and/or AES) which we defined as combined allergies (CA) group and controls without reported allergy symptoms, followed by separate analyses of AR and AES. We hypothesized that allergic symptoms would be associated with altered gut microbial diversity and functional potential, with both shared and symptom-specific microbial signatures across allergic phenotypes.

## Methods

### Sample characteristics from the FINRISK 2002 cohort microbiome study

The FINRISK 2002 study is a population-based survey of Finnish adults aged 25-74 years, based on a stratified random sample from multiple regions of Finland. Detailed study design and sampling procedures have been described previously^20,21^ and are provided in the **Supplementary Methods**. Participants completed health questionnaires, underwent clinical examinations, and provided stool samples for metagenomic sequencing. Allergic symptoms were assessed using two questionnaire items: “Have you ever had hay fever or other allergic nasal symptoms?” and “Have you ever had allergic eye symptoms?”. Response options were “no,” “yes, within the past 12 months,” or “yes, over a year ago.” Individuals reporting either affirmative response were classified as having allergic rhinitis (AR) or allergic eye symptoms (AES), respectively, representing lifetime symptom prevalence. To account for the substantial overlap between these conditions and the self-reported nature of the data, we additionally defined a combined allergies (CA) group comprising individuals reporting AR and/or AES. Controls were individuals reporting neither symptom.

The FINRISK 2002 protocol was approved by the Coordinating Ethical Committee of the Helsinki and Uusimaa Hospital District (558/E3/2001), and all participants provided written informed consent. Stool samples were collected, stored frozen, and subsequently subjected to shotgun metagenomic sequencing as previously described^21^. After quality control and exclusion criteria (**Supplementary Methods**), 5,595 individuals were included in the analyses, of whom 5,499 and 5,477 provided information on AR and AES, respectively.

### Taxonomic and functional profiling using shotgun metagenomics

Shotgun metagenomic sequences were quality filtered and adapter trimmed using Atropos^22^. Host-derived reads were removed by mapping against the human reference genome (GRCh38) using Bowtie2^23^. Filtered reads were taxonomically profiled using the Greengenes2 reference database (release 2022.10)^24^ and processed with the Woltka pipeline^25^. Taxonomic annotations were assigned using the QIIME2 q2-greengenes2 plugin QIIME2^26^. Functional profiling was performed using HUMAnN3^27^. UniRef90 gene families were aggregated into MetaCyc pathways and KEGG Orthologs (KOs) for downstream analyses. Detailed descriptions of the pipeline were provided in the **Supplementary Methods**. The computing resources of the Finnish IT Center for Science (CSC) were used for metagenome preprocessing. In differential abundance analysis, we only considered taxonomic groups detected with ≥ 1% prevalence at the ≥ 0.1% detection limit. This included 12 Phyla, 16 Classes, 34 Orders, 63 Families, 243 Genera, and 377 Species. Similarly, we analyzed predicted functions detected with ≥ 10% prevalence, retaining 166 MetaCyc pathways and 1,171 KOs.

### Statistical Analysis

The downstream statistical analysis was performed in R (version 4.3.1) using the TreeSummarizedExperiment (version 2.20.0) ^28^ data container with key R/Bioconductor packages, including mia^29^ and miaViz (versions 1.20.0). Alpha diversity was assessed using observed richness, Shannon diversity, and Faith’s phylogenetic diversity at the species level. Differences between allergic groups and controls were evaluated using the Wilcoxon rank-sum test, with p-value < 0.05 considered significant. Beta diversity was assessed using UniFrac dissimilarity and visualized by principal coordinates analysis (PCoA). Differences in overall community composition between groups were evaluated using permutational multivariate analysis of variance (PERMANOVA). The read count variation was controlled using niter parameter for both alpha and beta diversity.

Differential abundance analyses (DAA) of microbial taxa and predicted functions were performed using MaAsLin2 (version 1.26.0)^30^ as this provides implementation of linear models that showed favorable performance in recent DAA benchmarks^31^. We qualitatively analysed and visualized the enrichment patterns of the pathways using Enteropathway^32^. The analyses were adjusted for possible confounding factors such as age, BMI, sex, smoking status, and the geographical region in Finland (East and West). Models were adjusted for age, sex, body mass index, smoking status, and geographic region. Multiple testing was controlled using the Holm-Bonferroni correction, with adjusted p-values < 0.05 considered statistically significant.

To assess the robustness of associations across geographic regions, taxonomic and functional analyses were repeated separately in Eastern and Western Finland. Additional details on data processing, diversity calculations, and regional analyses are provided in the **Supplementary Methods**.

## Results

### Participants’ characteristics and microbial community composition

The characteristics of the study participants are reported in **Table 1**. Individuals with both AR and AES are statistically different in terms of age, BMI, smoking history, and sex compared to those without symptoms (p < 0.05). 1,199 individuals reported both allergy symptoms, and 2,305 individuals had either or both symptoms. Throughout the analyses, individuals reporting either AR and/or AES are collectively referred to as the CA group.

**Table 1.** Overview of the FINRISK 2002 cohort. . Data are presented as mean (SD) for continuous and n (%) for categorical variables. *The Mann-Whitney U test was used for continous variables; Fisher’s exact test was used for categorical variables.

| Group | All | Controls | Allergic Rhinitis |  | Allergic Eye Symptoms |  | Combined Allergies |  |
| --- | --- | --- | --- | --- | --- | --- | --- | --- |
| Variables |  |  | AR | p-value* | AES | p-value* | CA | p-value* |
| Sample size (n) | 5,595 | 3,175 (56.7%) | 1,950 (34.9%) | NA | 1,554 (27.8%) | NA | 2,305(41.2%) | NA |
| Baseline Age (years) | 49.6 (12.9) | 50.4 (12.6) | 48.2 (13.1) | 1.2e-08 | 47.1 (13.0) | 6.3e-16 | 48.3 (13.0) | 1.2e-08 |
| Body Mass Index (kg/m2) | 27.0 (4.7) | 27.0 (4.6) | 26.8 (4.7) | 0.055 | 26.9 (4.8) | 0.16 | 26.9 (4.7) | 0.17 |
| Number of smoking packs/year | 3.9 (10.5) | 4.4 (11.2) | 3.3 (9.3) | 0.0042 | 3.0 (8.6) | 5.5e-04 | 3.2 (9.2) | 0.001 |
| Male | 2646 (47.3%) | 1627 (51.2%) | 831 (42.6%) | 2.1e-09 | 609 (39.2%) | 6.4e-15 | 972 (42.2%) | 3.2e-11 |
| Current smokers | 1315 (23.5%) | 795 (25%) | 423 (21.7%) | 0.0056 | 319 (20.5%) | 5.2e-04 | 493 (21.4%) | 0.0016 |
| Living in Eastern Finland | 3,912 (69.9%) | 2,263 (71.3%) | 1,316 (67.5%) | 0.0043 | 1,016 (65.4%) | 4.2e-05 | 1,548 (67.2%) | 0.0012 |

There was no practical difference in overall species-level community composition between the CA group and controls, although the difference reached statistical significance (**Figure 1A**, PERMANOVA: R^2^=0.00096, p = 0.011). The observed species richness was significantly lower in AR, AES, and CA groups (**Figure 1B**, p = 0.0049; p = 3.9e-5; p = 0.0024, respectively). For the alpha diversity analyses, we detected differing results between Shannon and Faith indices; the Shannon diversity index did not capture any differences between allergy and control groups (**Figure 1C**, p > 0.05), whereas phylogenetically informed Faith index detected significant differences in all groups AR, AES, and CA (**Figure 1D**, p = 0.0049; p = 0.0001; p = 0.0027, respectively) suggesting phylogenetically different community diversities.

**Figure 1.**
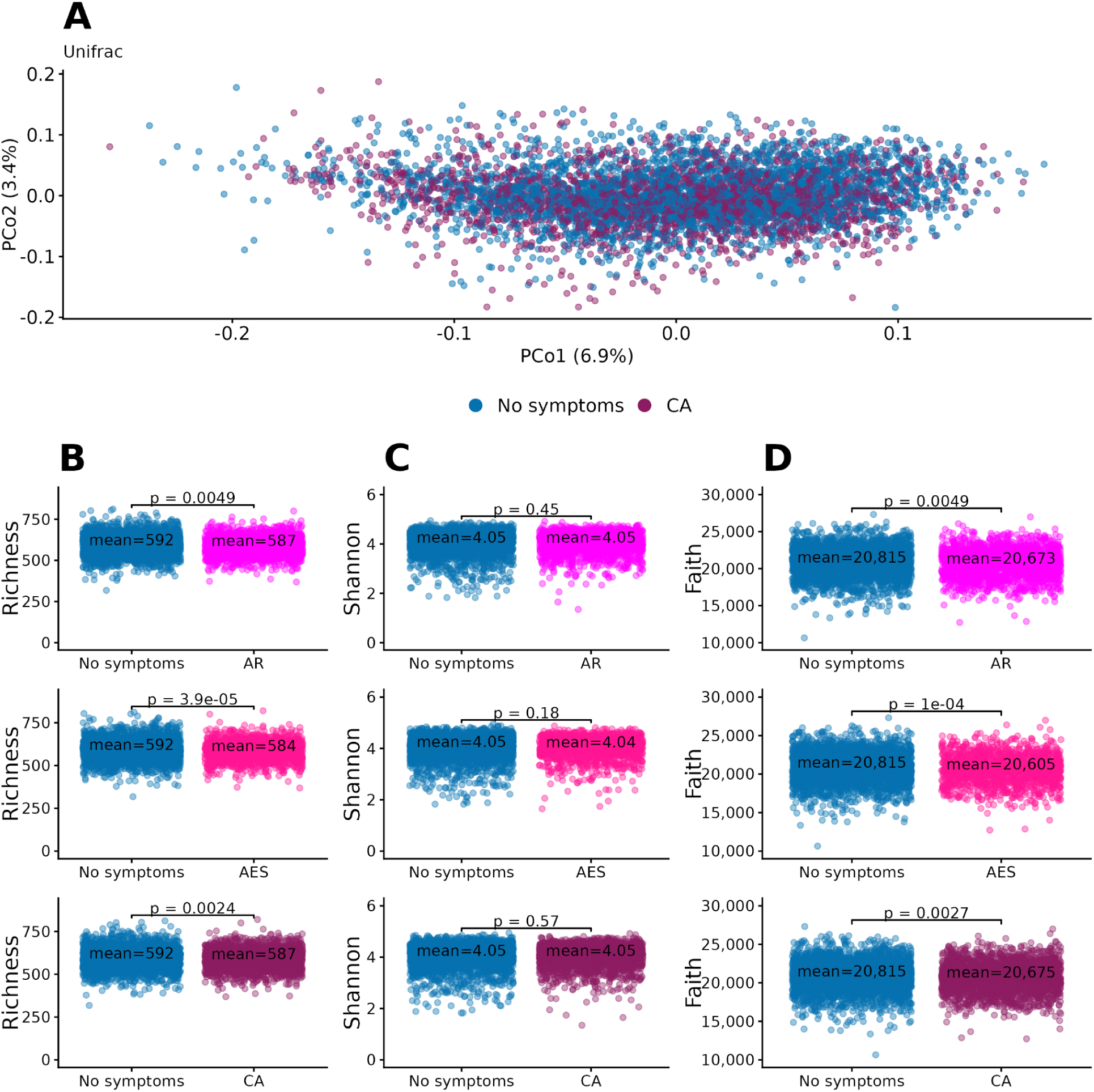
The overview of the taxonomic compositions and diversity of gut microbiota in individuals reporting allergy symptoms and in the control group. **A** Population variation based on microbial community composition as visualized by PCoA. **B** Observed richness in individuals with AR, AES, and CA, respectively, and the control group. **C** Shannon diversity index in individuals with AR, AES and CA, respectively, and the control group. **D** Faith phylogenetic diversity in individuals with AR, AES, and CA, respectively, and the control group. The indicated p-values were obtained with the Wilcoxon rank-sum test. AR stands for allergic rhinitis, AES for allergic eye symptoms, and the CA group is the union of both them.

### Enrichment of taxonomic groups associated with allergic symptoms

Overall, 34, 44 and 37 species were differentially abundant in AR, AES and CA groups, respectively, compared to the control group (**Table S1**; p < 0.05). Among these, 17 species, predominantly from the *Clostridia* class, were significantly enriched in all groups compared to the control group (**Figure 2**). *Sellimonas intestinalis*, *Ventrisoma faecale*, *Butyricicoccus_A 7730 sp900604335* taxa were also associated with asthma and chronic obstructive pulmonary disease (COPD) in the same cohort^33^. Furthermore, we also observed the enrichment of *Eggerthella lenta* in all groups, which have been reported to be more abundant in atopic dermatitis ^34^ and asthma^35^, related diseases to allergy. Moreover, *Massilioclostridium methylpentosum* and *Anaerobutyricum faecale* showed significant enrichment in AES versus controls only, with no corresponding enrichment observed in AR or CA group. On the other hand, 14 species were found to be less abundant in all allergic groups when compared to the control group, predominantly from the *Bacteroidales* order (*Prevotella*, *Cryptobacteroides*) along with several species from Actinobacteroidota phyla (*Collinsella* genus and *Slackia_A isoflavoniconvertens, Ellagibacter isourolithinifaciens*). While two species from Firmicutes_D (*Longicatena caecimuris* and *Fimiplasma intestinipullorum*) were enriched in the allergy groups, two species from Firmicutes_C phyla (*Megasphaera_A 38685 elsdenii* and *Phascolarctobacterium_A succinatutens*) were enriched in the control group.

**Figure 2.**
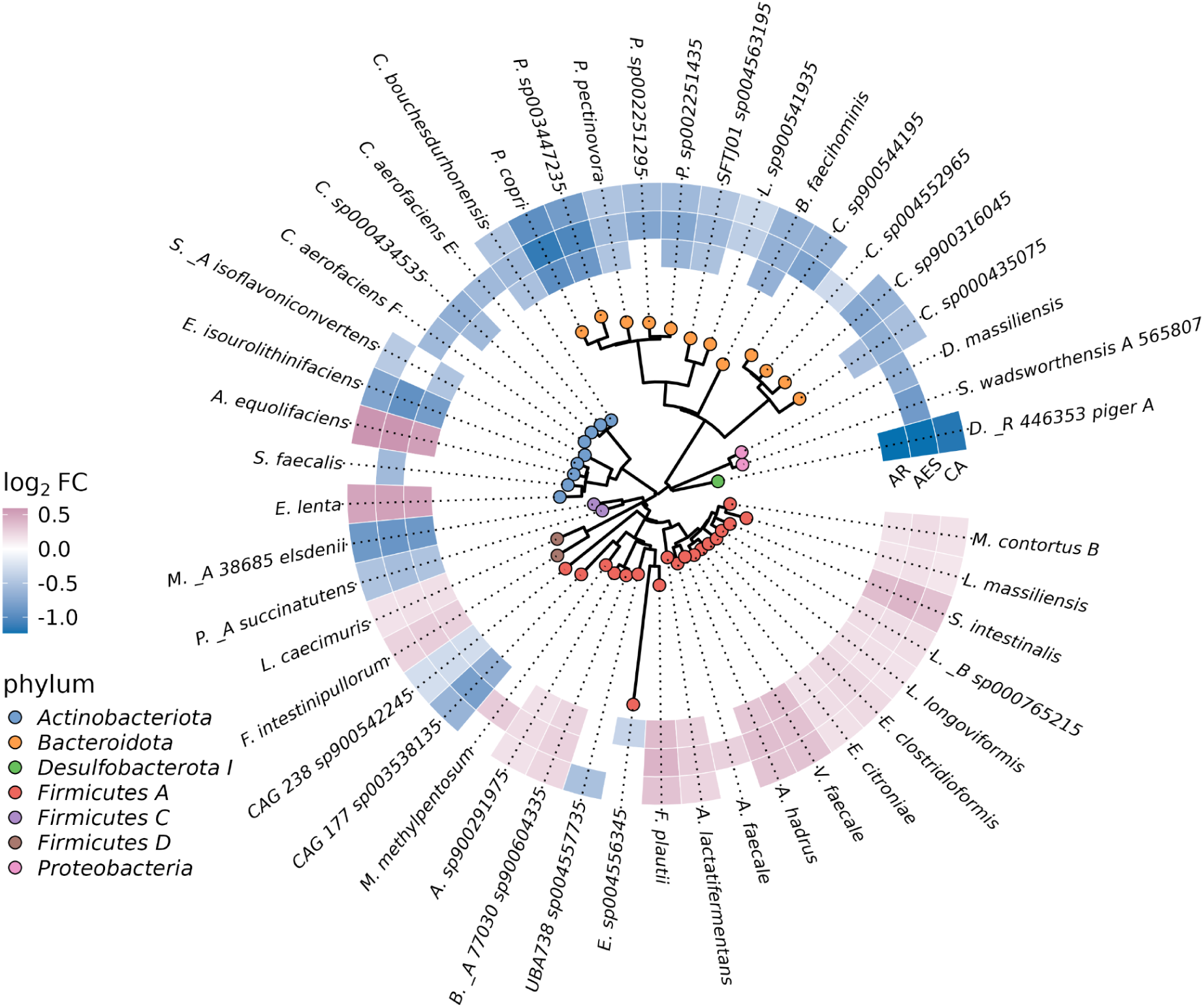
Estimated differences in species abundance in AR, AES, and CA compared with the control group presented along the phylogenetic tree. Heatmap showing log_2_(fold change) of MaAsLin2 coefficient representing the effect size of the allergic variables on microbial species based on relative abundance of species comparing AR, AES, and CA with controls. Pink indicates enrichment in the allergy groups, whereas blue indicates enrichment in the control group, and white indicates similar enrichment profiles between the groups as well as non-significant results. Only the significantly differentially abundant species in any of the groups are represented. The taxa predominantly in the *Clostridia* class (*Firmicutes A*) are more abundant in allergic groups, whereas the taxa in the *Prevotella* and *Cryptobacteroides* (*Bactereoidota*) less abundant.

### Distinct functional profiles in individuals with allergic symptoms

We aggregated the Uniref90-based HUMAnN3 predicted genes into KO functional groups (**Table S2**) and MetaCyc pathways (**Table S3**) and associated their relative abundances with the allergy symptoms (**Figure 3**). We used both profilings as they provide complementary insights into microbial function. Functional analyses revealed that most KOs and pathways were enriched in allergic individuals, whereas relatively few were enriched in controls, resulting in a one-sided enrichment pattern in the volcano plots (**Figure S1**). While this pattern may reflect coordinated shifts in microbial metabolic potential associated with allergic conditions, it may also partly arise from technical and statistical properties of metagenomic functional profiling, including the compositional nature of relative abundance data, sparse or low-abundance KO profiles and pathway redundancy^36^. Nevertheless, several enriched functions highlighted biologically plausible mechanisms linking gut microbial metabolism to allergic symptoms as described below.

**Figure 3.**
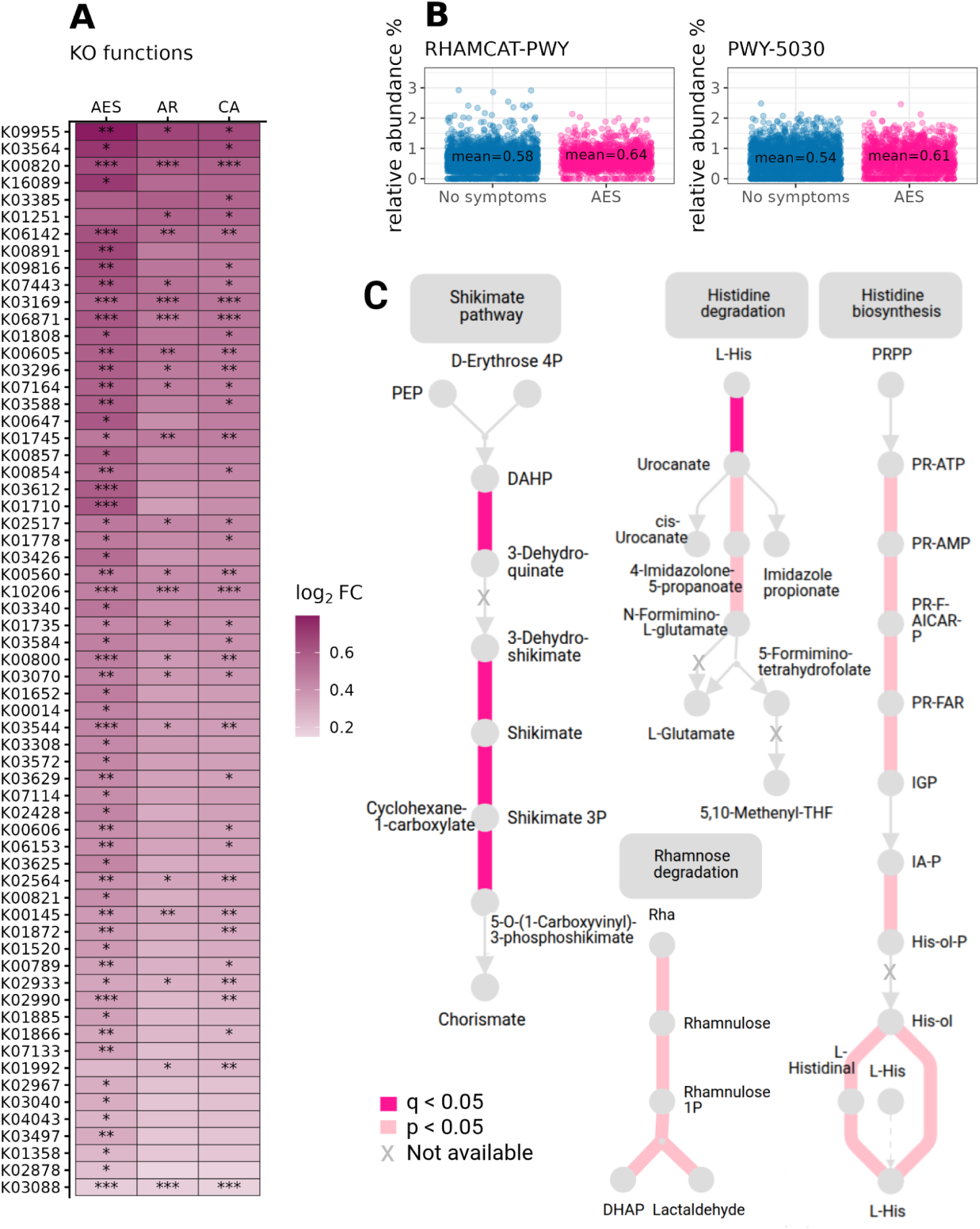
Differentially abundant KO and Metacyc pathway functional features in AR, AES and Allergies. **A** Heatmap illustrates the differential abundance of KOs in individuals with AR, AES and CA compared to the control group (with MaAsLin2). All significant KOs are enriched in allergic groups compared to control, while AES captures the strongest associations among many KOs. **B** Relative abundances (%) of significant (q < 0.05) Metacyc pathways, l-rhamnose (RHAMCAT-PWY) and L-histidine degradation III (PWY.5030), between the AES and control groups. **C** Graphical illustration of selected KOs (Shikimate, Rhamnose degradation and Histidine pathways) using Enteropathway tool. Represent consistent enrichment patterns among allergic individuals, where Shikimate pathways are statistically significantly enriched (q < 0.05), while others do not reach significance after p-value correction.

In the KO-based analysis, 38 functions were significantly enriched in the CA group, whereas no functions were significantly enriched in controls (**Figure 3A**). Among these, 21 KOs were consistently enriched across the CA, AR, and AES analyses. The AES group exhibited a larger set of functions enriched compared with controls, including 14 shared with the CA group but not observed in AR and 25 uniquely enriched in AES. Notably, the effect sizes of associations were consistently higher in the AES group across both shared and AR-specific KOs.

Shared enrichments across allergy groups included KOs involved in amino sugar and aminocyclitol metabolism (K00820, K02564), shikimate-related pathways (K01735, K00800), and amino acid metabolism, including L-histidine degradation (K01745) and L-ornithine biosynthesis (K00145, K00821) (**Figure 3C**). AES further complemented these pathways through enrichment of additional shikimate-related KOs (K00014, K00891) and amino acid metabolism functions. Consistent with the KO-level findings, MetaCyc pathway analysis identified enrichment of L-histidine degradation III (PWY.5030) and L-rhamnose catabolism (RHAMCAT-PWY), the latter uniquely enriched in AES (**Figure 3B,C**)

Many AES-associated KOs belonged to broader functional categories already enriched in the CA group, particularly amino acid, carbohydrate, and nucleotide metabolism. These included functions related to branched-chain amino acid biosynthesis (K01652), lysine biosynthesis (K00821), methionine metabolism (K00789), pyrimidine metabolism (K00857, K01520), amino sugar degradation (K01788, K02564), and ketone and keto-acid metabolism (K01652, K03612). Together, these results highlight shifts in both metabolic activity and immune-related functional pathways in individuals with allergic symptoms.

### Replication of taxonomic and functional association patterns across regions

AR was reported by 33.6% (n = 1,316) and 37.7% (n = 634) of participants in Eastern and Western Finland, respectively, while AES was reported by 26.0% (n = 1,016) and 31.7% (n = 538). Although overall taxonomic association patterns were similar across the full dataset and regional subsets, no species were consistently significant across all regions (**Figure S2A**, **Table S4**, **Table S5**).

Several members of the *Clostridia* class, *Fimiplasma intestinipullorum*, and *Adlercreutzia equolifaciens* were significantly enriched in the allergy groups in the overall dataset and in the Eastern Finland subset. While these species also showed a similar enrichment trend in the AR group in western Finland, the association was not significant (q > 0.05). Likewise, several Bacteroidales and Prevotella-related taxa were consistently depleted in allergic groups, although the specific associations reaching significance varied by region. Notably, species that were significant in one region generally showed similar effect directions in the other region. Consistent with previous studies in the same cohort^37^, these findings suggest that microbial association patterns are largely reproducible across regions, although statistical significance varies.

Functional analyses showed a similar pattern (**Figure S2B**, **Table S6**, **Table S7**). While few functional features were consistently statistically significant across regions, the overall enrichment and depletion trends were largely concordant between the full dataset and regional subsets. Only K06871, an uncharacterized protein, was significantly enriched across multiple allergic symptom groups in both the overall dataset and regional analyses. No MetaCyc pathways reached statistical significance (**Table S8**, **Table S9**) in the regional subsets, although the same directional trends were observed. Together, these findings indicate that regional variation influences the strength of statistical evidence rather than the overall pattern of taxonomic and functional associations.

## Discussion

AR and AES are closely related allergic conditions that share common immune mechanisms with asthma, atopic dermatitis, and food allergy^38^. Although early-life gut microbiota has been implicated in allergic disease development^17,38^, evidence from adult populations remains limited. In this large population-based study of Finnish adults, individuals with allergic symptoms exhibited distinct gut microbial taxonomic and functional profiles. Given the substantial overlap and shared pathophysiology between AR and AES, including interconnected nasal-ocular inflammatory pathways and neurogenic responses affecting both mucosal surfaces^3^, we additionally analyzed a broader allergy group comprising individuals reporting AR and/or AES symptoms (CA). This combined grouping may better capture the overall allergic phenotype in self-reported data. The largely overlapping microbiome signatures observed across AR, AES, and CA support a shared allergic etiology, with associated taxa previously linked to related allergic and respiratory diseases and functional enrichments involving mucosal carbohydrate processing, histidine metabolism, and other immune-relevant pathways. The taxonomic and functional findings of the current study align with research findings linking gut microbiome dysbiosis to allergic conditions^7,8^. Interestingly, despite similar sample sizes, AES exhibited stronger effect sizes and additional functional associations compared with AR and CA groups. This may suggest that AES represents a more homogeneous or specific allergic phenotype, whereas AR may include greater clinical heterogeneity, including non-allergic rhinitis presentations^39^, potentially diluting microbiome-associated signals.

High community diversity has been suggested to be beneficial for a balanced immune response^13,14^. In line with this, we observed significantly higher observed richness and Faith’s phylogenetic diversity in controls compared with the CA, AR, and AES groups. However, we did not observe significant difference in Shannon diversity (**Figure 1 B,C,D**). Phylogeny-aware metrics such as Faith’s phylogenetic diversity incorporate evolutionary relationships among taxa through branch lengths in a phylogenetic tree and may therefore provide more informative comparisons of microbial community structure across taxonomic scales^40^. The significant differences observed in Faith’s index but not in Shannon diversity may suggest that allergic symptoms are associated more strongly with a loss of phylogenetically distinct microbial lineages than with changes in overall richness or evenness or species abundance. Notably, the allergy groups were enriched with taxa that have been linked to respiratory diseases (**Figure 2**). For example, the class *Clostridia* and *E. lenta* have been positively linked to the risk of asthma^33,41^ and COPD^33^. In our data, a higher percentage of individuals who reported both allergic symptoms developed asthma during the 20 years of follow-up period compared to the control group (**Figure S3**). These\ observations suggest potential links between gut microbiome composition and systemic immune responses impacting respiratory health.

In line with the taxonomic analysis, we observed shared enrichment of functional features in all allergy groups, which suggests common functional adaptations linked to allergic responses. We observed enrichment of KOs linked to the processing of mucosal carbohydrates which contribute to the intestinal barrier integrity and immune regulation^42^. Increased microbial degradation of mucosal glycans has previously been linked to impaired mucus barrier function and enhanced susceptibility to allergic sensitization in the gut^42,43^. In the context of AR and AES, such alterations may contribute to heightened mucosal inflammatory responses through increased exposure to microbial and environmental antigens, supporting the concept of interconnected gut-mucosal immune axes, including the gut-nose^44^ and gut-eye axis^45^. We also observed enrichment of shikimate-related pathways in all allergy groups (**Figure 3C**), which are absent in mammalian cells and are involved in the microbial biosynthesis of aromatic amino acids such as phenylalanine, tyrosine, and tryptophan. Although metabolites derived from these pathways have been implicated in immune regulation, including modulation of IgE-mediated mast cell responses, promotion of regulatory T-cell differentiation, and attenuation of allergic inflammation^8^, the observed enrichment at the functional gene level does not necessarily translate into increased production of immunoregulatory metabolites in the host. This is supported by evidence that tryptophan metabolism is tightly regulated in allergic disease through immune pathways such as interferon-γ–induced indoleamine 2,3-dioxygenase-1 (IDO-1) activity, with altered tryptophan levels reported in patients with allergic rhinitis in a context-dependent manner^46^.

More broadly, we observed allergy-associated enrichment in amino acid metabolism pathways, including valine, leucine, isoleucine, ornithine, and histidine metabolism. These pathways have previously been linked to TH2-dominant immune responses, airway hyperresponsiveness, and allergic inflammation^47^. Importantly, we detected concurrent enrichment of histidine biosynthesis (**Figure 3C**) and degradation pathways suggesting enhanced microbial turnover of histidine within the gut ecosystem in allergic individuals. This may reflect increased metabolic demand or cross-feeding interactions among microbial taxa, rather than a unidirectional shift in histidine availability. Given that histidine is a precursor of histamine, a key mediator of allergic responses^8,48^, these findings point to potential alterations in amino acid metabolism in allergic conditions, although the functional consequences for host histamine levels remain to be determined.

Interestingly, AES exhibited broader functional enrichment patterns and generally stronger effect sizes compared with AR, including AES-specific enrichment of ketone and keto-acid metabolism pathways. This may indicate that AES captures a more pronounced or metabolically active allergic phenotype, potentially providing a broader representation of allergic mucosal inflammation together with AR. At the same time, these AES-associated signatures may also partly reflect treatment-related effects, as individuals with ocular symptoms are more likely to use antihistamines or ophthalmic anti-allergy medications that could influence gut microbial metabolism. Ketone-related chemical structures are present in several anti-allergic agents, including ketotifen fumarate^5^, although the microbial pathways identified here are not directly equivalent to these compounds.

We identified several taxa that were depleted in allergic individuals, including multiple *Prevotella* species. This is consistent with previous studies reporting an inverse association between *Prevotella* abundance and AR, potentially reflecting the importance of early microbial exposures for immune development^14^. *Prevotella* and other depleted taxa, such as *Megasphaera elsdenii*, have been linked to the production of short-chain fatty acids (SCFAs), microbial metabolites that promote immune tolerance and anti-inflammatory responses^49–53^. Reduced abundance of these taxa may therefore indicate diminished microbial capacity to support immune regulation in allergic individuals. More broadly, the depletion of *Prevotella* and *Cryptobacteroides* is consistent with observations that these taxa are more prevalent in non-industrialized populations^54,55^. These findings underscore the potential roles of exposure to diverse microbes in modulating immune functions, which may alleviate the risk of developing allergic conditions.

Our analysis revealed consistent trends in microbial species and functional features associated with allergy symptoms across the entire dataset, as well as in the regional subsets. However, the absence of statistical significance in some cases is likely due to variations in sample size and statistical power. While the regional dataset is not independent, the replication of microbial patterns within this genetically distinct subpopulation supports the idea that certain taxa are consistently associated with allergic conditions across different Finnish regions. However, the region-specific statistically significant enrichment also highlights the possibility of localized influences, for example, dietary habits, environmental exposure, or access to healthcare may vary between regions and could influence the composition of the gut microbiota. These findings underscore the importance of accounting for geographic variation in gut microbiome studies^56^, as both consistent cross-regional signals and region-specific associations were observed.

This study should be interpreted in light of several limitations. First, allergy status was based on self-reported symptoms, which may introduce recall bias and misclassification. Second, the study included only adults. As AR usually develops early in life, follow-up from childhood would be essential to more accurately assess the role of the gut microbiota in disease development. The substantial overlap between AR and AES in our cohort (n = 1,199), consistent with previous reports^57^, may also contribute to their shared microbial signatures. Furthermore, the cross-sectional design precludes causal inference, and future longitudinal studies are required to determine whether microbiome alterations precede or result from allergic symptoms. Finally, the use of allergy-related medications, including antihistamines and corticosteroid nasal sprays, may have influenced gut microbial composition^58^ and therefore contributed to some of the observed associations.

Although the cross-sectional and self-reported nature of the data limits causal interpretation, our findings support the relevance of gut microbial metabolic potential in allergic conditions and emphasize the importance of considering phenotypic heterogeneity and regional structure in microbiome studies. Notably, by focusing on an adult population with a mature and environmentally shaped gut microbiome, this study complements the predominantly pediatric literature and provides insights into allergic disease mechanisms beyond early-life immune development windows. Collectively, this work provides a basis for future mechanistic and longitudinal studies aimed at disentangling shared and disease-specific microbial pathways in allergic disease.

## Declarations

### Ethics approval and consent to participate

The study protocol for FINRISK 2002 was approved by the Coordinating Ethics Committee of the Helsinki University Hospital District (Helsinki, Finland) with reference number 558/E3/2001, and all participants provided written informed consent.

### Consent for publication

Not applicable.

### Availability of data and material

The metagenomic data from FINRISK 2002 samples are available from the European Genome-Phenome Archive (accession number EGAD00001007035). The data supporting the findings of this study are not openly available due to sensitive health information of individuals. Access to the data is available through the THL Biobank following the standard application procedure (https://thl.fi/en/research-and-development/thl-biobank/for-researchers/application-process). For further verification, THL Biobank can be contacted at. The source code for the analysis is available (https://github.com/finrisk2002/article-2026-allergic-rhinitis-and-eye-symptoms).

### Authors’ contributions

H.L., P.P.E., V.V., L.L. conceived of the study, designed the study, analyzed the data, and wrote the manuscript. N.B., G.Mu, K.P., and M.R contributed to data analysis. G.Me., P.J., R.K., A.S.H., V.S., T.N. and L.L. provided intellectual input on study goals and design and contributed to manuscript preparation.

R.K. acquired shotgun metagenomics data. V.S. and T.N. contributed to recruitment of study participants and the collection of biomaterials and clinical data. All authors reviewed, edited and approved the final version of the manuscript.

## Supporting information

Supplementary Methods, Supplementary Results

## Acknowledgements

The authors thank all participants in the FINRISK 2002 study and Tara Schwartz for assistance with laboratory work, and wish to acknowledge CSC - IT Center for Science, Finland, for computational resources.

## Abbreviations

AR: Allergic Rhinitis
AES: Allergic Eye Symptoms
CA: Combined Allergies
TH2: T-helper type 2
GA²LEN: Global Allergy and Asthma Excellence Network
PERMANOVA: Permutational Multivariate Analysis of Variance
GG2: Greengenes2
WoL: Web of Life
GTDB: Genome Taxonomy Database
KEGG: Kyoto Encyclopedia of Genes and Genomes
KO: KEGG Orthologs
QIIME: Quantitative Insights Into Microbial Ecology
PCoA: Principal coordinates analyses
MaAsLin2: Microbiome Multivariable Associations with Linear Models 2

## References

1. Jousilahti P, Haahtela T, Laatikainen T, Mäkelä M, Vartiainen E. Asthma and respiratory allergy prevalence is still increasing among Finnish young adults. Eur Respir J. 2016 Mar 1;47(3):985–7. doi:10.1183/13993003.01702-2015 PubMed PMID: 26677942.

2. Reijula J, Latvala J, Mäkelä M, Siitonen S, Saario M, Haahtela T. Long-term trends of asthma, allergic rhinitis and atopic eczema in young Finnish men: a retrospective analysis, 1926-2017. Eur Respir J. 2020 Dec;56(6):1902144. doi:10.1183/13993003.02144-2019 PubMed PMID: 32764114.

3. Spicer S, Davis A, Ellis AK. When the eyes and nose collide: The shared pathways of rhinoconjunctivitis. Ann Allergy Asthma Immunol. 2026 Apr 5. doi:10.1016/j.anai.2026.04.002

4. Larsson ML, Loit HM, Meren M, Põlluste J, Magnusson A, Larsson K, et al. Passive smoking and respiratory symptoms in the FinEsS Study. Eur Respir J. 2003 Apr 1;21(4):672–6. doi:10.1183/09031936.03.00033702 PubMed PMID: 12762355.

5. Bielory L. Allergic Conjunctivitis and the Impact of Allergic Rhinitis. Curr Allergy Asthma Rep. 2010 Mar 1;10(2):122–34. doi:10.1007/s11882-010-0087-1

6. Savouré M, Bousquet J, Leynaert B, Renuy A, Siroux V, Goldberg M, et al. Rhinitis phenotypes and multimorbidities in the general population: the CONSTANCES cohort. Eur Respir J. 2023 Feb 9;61(2). doi:10.1183/13993003.00943-2022 PubMed PMID: 36202419.

7. Aguilera AC, Dagher IA, Kloepfer KM. Role of the Microbiome in Allergic Disease Development. Curr Allergy Asthma Rep. 2020 Jun 16;20(9):44. doi:10.1007/s11882-020-00944-2 PubMed PMID: 32548788; PubMed Central PMCID: PMC7702839.

8. Losol P, Wolska M, Wypych TP, Yao L, O’Mahony L, Sokolowska M. A cross talk between microbial metabolites and host immunity: Its relevance for allergic diseases. Clin Transl Allergy. 2024;14(2):e12339. doi:10.1002/clt2.12339

9. Joos R, Boucher K, Lavelle A, Arumugam M, Blaser MJ, Claesson MJ, et al. Examining the healthy human microbiome concept. Nat Rev Microbiol. 2025 Mar;23(3):192–205. doi:10.1038/s41579-024-01107-0

10. Murdaca G, Greco M, Borro M, Gangemi S. Hygiene hypothesis and autoimmune diseases: A narrative review of clinical evidences and mechanisms. Autoimmun Rev. 2021 Jul;20(7):102845. doi:10.1016/j.autrev.2021.102845 PubMed PMID: 33971339.

11. Haahtela T, Alenius H, Auvinen P, Fyhrquist N, von Hertzen L, Jousilahti P, et al. A short history from Karelia study to biodiversity and public health interventions. Front Allergy. 2023 Mar 14;4. doi:10.3389/falgy.2023.1152927

12. Ruokolainen L, Fyhrquist N, Laatikainen T, Auvinen P, Fortino V, Scala G, et al. Immune-microbiota interaction in Finnish and Russian Karelia young people with high and low allergy prevalence. Clin Exp Allergy. 2020;50(10):1148–58. doi:10.1111/cea.13728

13. Watts AM, West NP, Zhang P, Smith PK, Cripps AW, Cox AJ. The Gut Microbiome of Adults with Allergic Rhinitis Is Characterised by Reduced Diversity and an Altered Abundance of Key Microbial Taxa Compared to Controls. Int Arch Allergy Immunol. 2021;182(2):94–105. doi:10.1159/000510536 PubMed PMID: 32971520.

14. Sahoyama Y, Hamazato F, Shiozawa M, Nakagawa T, Suda W, Ogata Y, et al. Multiple nutritional and gut microbial factors associated with allergic rhinitis: the Hitachi Health Study. Sci Rep. 2022 Mar 1;12(1):3359. doi:10.1038/s41598-022-07398-8

15. Hua X, Goedert JJ, Pu A, Yu G, Shi J. Allergy associations with the adult fecal microbiota: Analysis of the American Gut Project. eBioMedicine. 2016 Jan 1;3:172–9. doi:10.1016/j.ebiom.2015.11.038 PubMed PMID: 26870828.

16. Liu X, Tao J, Li J, Cao X, Li Y, Gao X, et al. Dysbiosis of Fecal Microbiota in Allergic Rhinitis Patients. Am J Rhinol Allergy. 2020 Sep 1;34(5):650–60. doi:10.1177/1945892420920477

17. Kallio S, Jian C, Korpela K, Kukkonen AK, Salonen A, Savilahti E, et al. Early-life gut microbiota associates with allergic rhinitis during 13-year follow-up in a Finnish probiotic intervention cohort. Microbiol Spectr. 2024 Apr 30;12(6):e04135–23. doi:10.1128/spectrum.04135-23

18. Liu K, Cai Y, Song K, Yuan R, Zou J. Clarifying the effect of gut microbiota on allergic conjunctivitis risk is instrumental for predictive, preventive, and personalized medicine: a Mendelian randomization analysis. EPMA J. 2023 Jun 1;14(2):235–48. doi:10.1007/s13167-023-00321-9

19. Hua X, Goedert JJ, Pu A, Yu G, Shi J. Allergy associations with the adult fecal microbiota: Analysis of the American Gut Project. EBioMedicine. 2016 Jan;3:172–9. doi:10.1016/j.ebiom.2015.11.038 PubMed PMID: 26870828; PubMed Central PMCID: PMC4739432.

20. Borodulin K, Tolonen H, Jousilahti P, Jula A, Juolevi A, Koskinen S, et al. Cohort Profile: The National FINRISK Study. Int J Epidemiol. 2018 Jun 1;47(3):696–696i. doi:10.1093/ije/dyx239

21. Salosensaari A, Laitinen V, Havulinna AS, Meric G, Cheng S, Perola M, et al. Taxonomic Signatures of Long-Term Mortality Risk in Human Gut Microbiota. medRxiv. 2020 Jan 13;2019.12.30.19015842. doi:10.1101/2019.12.30.19015842

22. Didion JP, Martin M, Collins FS. Atropos: specific, sensitive, and speedy trimming of sequencing reads. PeerJ. 2017 Aug 30;5:e3720. doi:10.7717/peerj.3720 PubMed PMID: 28875074; PubMed Central PMCID: PMC5581536.

23. Langmead B, Salzberg SL. Fast gapped-read alignment with Bowtie 2. Nat Methods. 2012 Apr;9(4):4. doi:10.1038/nmeth.1923

24. McDonald D, Jiang Y, Balaban M, Cantrell K, Zhu Q, Gonzalez A, et al. Greengenes2 unifies microbial data in a single reference tree. Nat Biotechnol. 2023 Jul 27;1–4. doi:10.1038/s41587-023-01845-1

25. Q Z, S H, A G, I M, D M, N H, et al. Phylogeny-Aware Analysis of Metagenome Community Ecology Based on Matched Reference Genomes while Bypassing Taxonomy. mSystems. 2022 Apr 26;7(2). doi:10.1128/msystems.00167-22 PubMed PMID: 35369727.

26. Bolyen E, Rideout JR, Dillon MR, Bokulich NA, Abnet CC, Al-Ghalith GA, et al. Reproducible, interactive, scalable and extensible microbiome data science using QIIME 2. Nat Biotechnol. 2019 Aug;37(8):852–7. doi:10.1038/s41587-019-0209-9 PubMed PMID: 31341288; PubMed Central PMCID: PMC7015180.

27. Beghini F, McIver LJ, Blanco-Míguez A, Dubois L, Asnicar F, Maharjan S, et al. Integrating taxonomic, functional, and strain-level profiling of diverse microbial communities with bioBakery 3. eLife. 10:e65088. doi:10.7554/eLife.65088 PubMed PMID: 33944776; PubMed Central PMCID: PMC8096432.

28. Huang R, Soneson C, Ernst FGM, Rue-Albrecht KC, Yu G, Hicks SC, et al. TreeSummarizedExperiment: a S4 class for data with hierarchical structure [Internet]. F1000Research; 2021 [cited 2024 Nov 11]. Available from: https://f1000research.com/articles/9-1246 doi:10.12688/f1000research.26669.2

29. Borman T, Benedetti G, Muluh G, Raulo A, Valderrama B, Sannikov A, et al. Orchestrating Microbiome Analysis with Bioconductor [Internet]. bioRxiv; 2025 [cited 2025 Dec 22]. p. 2025.10.29.685036. Available from: https://www.biorxiv.org/content/10.1101/2025.10.29.685036v1 doi:10.1101/2025.10.29.685036

30. Lin H, Eggesbø M, Peddada SD. Linear and nonlinear correlation estimators unveil undescribed taxa interactions in microbiome data. Nat Commun. 2022 Aug 23;13(1):4946. doi:10.1038/s41467-022-32243-x

31. Pelto J, Auranen K, Kujala JV, Lahti L. Elementary methods provide more replicable results in microbial differential abundance analysis. Brief Bioinform. 2025 Mar 4;26(2):bbaf130. doi:10.1093/bib/bbaf130

32. Shiroma H, Darzi Y, Terajima E, Nakagawa Z, Tsuchikura H, Tsukuda N, et al. Enteropathway: the metabolic pathway database for the human gut microbiota. Brief Bioinform. 2024 Sep 1;25(5):bbae419. doi:10.1093/bib/bbae419

33. Liu Y, Teo SM, Méric G, Tang HHF, Zhu Q, Sanders JG, et al. The gut microbiome is a significant risk factor for future chronic lung disease. J Allergy Clin Immunol. 2023 Apr 1;151(4):943–52. doi:10.1016/j.jaci.2022.12.810 PubMed PMID: 36587850.

34. Wang Y, Liu T, Wan Z, Wang L, Hou J, Shi M, et al. Investigating causal relationships between the gut microbiota and allergic diseases: A mendelian randomization study. Front Genet. 2023 Apr 12;14:1153847. doi:10.3389/fgene.2023.1153847 PubMed PMID: 37124612; PubMed Central PMCID: PMC10130909.

35. Wang Q, Li F, Liang B, Liang Y, Chen S, Mo X, et al. A metagenome-wide association study of gut microbiota in asthma in UK adults. BMC Microbiol. 2018 Sep 12;18(1):114. doi:10.1186/s12866-018-1257-x

36. Zielińska K, Pantiukh K, Org E, Łabaj PP, Kosciolek T. Moving from a taxonomic to a functional perspective in global microbiome analysis requires optimizing multiplexing ratios. mSystems. 2026 May 29;11(6):e00144–26. doi:10.1128/msystems.00144-26

37. Ruuskanen MO, Erawijantari PP, Havulinna AS, Liu Y, Méric G, Tuomilehto J, et al. Gut Microbiome Composition Is Predictive of Incident Type 2 Diabetes in a Population Cohort of 5,572 Finnish Adults. Diabetes Care. 2022 Jan 31;45(4):811–8. doi:10.2337/dc21-2358

38. Wang J, Zhou Y, Zhang H, Hu L, Liu J, Wang L, et al. Pathogenesis of allergic diseases and implications for therapeutic interventions. Signal Transduct Target Ther. 2023 Mar 24;8(1):1–30. doi:10.1038/s41392-023-01344-4

39. Dykewicz MS, Wallace DV, Amrol DJ, Baroody FM, Bernstein JA, Craig TJ, et al. Rhinitis 2020: A practice parameter update. J Allergy Clin Immunol. 2020 Oct 1;146(4):721–67. doi:10.1016/j.jaci.2020.07.007 PubMed PMID: 32707227.

40. Bewick SA, Camper BT. Phylogenetic Measures of the Core Microbiome [Internet]. bioRxiv; 2025 [cited 2026 May 20]. p. 2024.10.10.617603. Available from: https://www.biorxiv.org/content/10.1101/2024.10.10.617603v2 doi:10.1101/2024.10.10.617603

41. Wang Z, Qu J, Chang C, Sun Y. Association of the gut microbiome and different phenotypes of COPD and asthma: a bidirectional Mendelian randomization study. Microbiol Spectr. 2024 Oct 7;12(11):e01760–24. doi:10.1128/spectrum.01760-24

42. Tiffany E, Kim KS, Sittipo P, Lee DW, Lee YK. Mucin-degrading gut bacteria: context-dependent roles in intestinal homeostasis and disease. Gut Microbes. 2026 Jan 23;18(1):2614054. doi:10.1080/19490976.2026.2614054 PubMed PMID: 41572837.

43. Parrish A, Boudaud M, Kuehn A, Ollert M, Desai MS. Intestinal mucus barrier: a missing piece of the puzzle in food allergy. Trends Mol Med. 2022 Jan 1;28(1):36–50. doi:10.1016/j.molmed.2021.10.004

44. Sun G, Zhao S, Huang H, Guan W, Wang X, Zhang H, et al. Integrated gut microbiome and metabolomics analysis reveals microbial-metabolic cross-talk in allergic rhinitis. Front Microbiol. 2025 Nov 12;16. doi:10.3389/fmicb.2025.1652915

45. Campagnoli LIM, Varesi A, Barbieri A, Marchesi N, Pascale A. Targeting the Gut–Eye Axis: An Emerging Strategy to Face Ocular Diseases. Int J Mol Sci. 2023 Aug 28;24(17):13338. doi:10.3390/ijms241713338 PubMed PMID: 37686143; PubMed Central PMCID: PMC10488056.

46. Gostner JM, Becker K, Kofler H, Strasser B, Fuchs D. Tryptophan Metabolism in Allergic Disorders. Int Arch Allergy Immunol. 2016;169(4):203–15. doi:10.1159/000445500 PubMed PMID: 27161289; PubMed Central PMCID: PMC5433561.

47. Liu QD, Pan GX, Yan YJ, Li JW, Zhang JJ, Liu HL, et al. Metabolomic profiles in allergic rhinitis: A systematic review and meta-analysis. Ann Allergy Asthma Immunol. 2025 Jan 15. doi:10.1016/j.anai.2024.12.022

48. Chun Y, Grishin A, Rose R, Zhao W, Arditi Z, Zhang L, et al. Longitudinal dynamics of the gut microbiome and metabolome in peanut allergy development. J Allergy Clin Immunol. 2023 Dec 1;152(6):1569–80. doi:10.1016/j.jaci.2023.08.012 PubMed PMID: 37619819.

49. Oka A, Kanai K, Higaki T, Makihara S, Noda Y, Kariya S, et al. Prevotella induces IL-10 production in monocytic cells. Rhinol Online. 2024 Mar 1;7(7):25–8. doi:10.4193/RHINOL/23.025

50. Vuillermin PJ, O’Hely M, Collier F, Allen KJ, Tang MLK, Harrison LC, et al. Maternal carriage of Prevotella during pregnancy associates with protection against food allergy in the offspring. Nat Commun. 2020 Mar 24;11(1):1452. doi:10.1038/s41467-020-14552-1

51. Shetty SA, Marathe NP, Lanjekar V, Ranade D, Shouche YS. Comparative genome analysis of Megasphaera sp. reveals niche specialization and its potential role in the human gut. PloS One. 2013 Jan 1;8(11):e79353. doi:10.1371/journal.pone.0079353 PubMed PMID: 24260205; PubMed Central PMCID: PMC3832451.

52. Sasaki M, Suaini NHA, Afghani J, Heye KN, O’Mahony L, Venter C, et al. Systematic review of the association between short-chain fatty acids and allergic diseases. Allergy. 2024;79(7):1789–811. doi:10.1111/all.16065

53. Borewicz K, Zhao Y, Zhu Y. Daily intake of a dairy-based nutritional supplement improved self-reported gastrointestinal symptoms and modulated microbiota in adult Chinese volunteers. Sci Rep. 2024 Nov 19;14(1):28651. doi:10.1038/s41598-024-79360-9

54. Rühlemann MC, Bang C, Gogarten JF, Hermes BM, Groussin M, Waschina S, et al. Functional host-specific adaptation of the intestinal microbiome in hominids. Nat Commun. 2024 Jan 6;15(1):326. doi:10.1038/s41467-023-44636-7

55. Singh H, Wiscovitch-Russo R, Kuelbs C, Espinoza J, Appel AE, Lyons RJ, et al. Multiomic Insights into Human Health: Gut Microbiomes of Hunter-Gatherer, Agropastoral, and Western Urban Populations. bioRxiv. 2024 Sep 4;2024.09.03.611095. doi:10.1101/2024.09.03.611095 PubMed PMID: 39282340; PubMed Central PMCID: PMC11398329.

56. He Y, Wu W, Zheng HM, Li P, McDonald D, Sheng HF, et al. Regional variation limits applications of healthy gut microbiome reference ranges and disease models. Nat Med. 2018 Oct;24(10):1532–5. doi:10.1038/s41591-018-0164-x

57. Hom MM, Bielory L. The anatomical and functional relationship between allergic conjunctivitis and allergic rhinitis. Allergy Rhinol. 2013;4(3):e110–9. doi:10.2500/ar.2013.4.0067 PubMed PMID: 24498515; PubMed Central PMCID: PMC3911799.

58. Vich Vila A, Collij V, Sanna S, Sinha T, Imhann F, Bourgonje AR, et al. Impact of commonly used drugs on the composition and metabolic function of the gut microbiota. Nat Commun. 2020 Jan 17;11(1):362. doi:10.1038/s41467-019-14177-z

