## Supplementary Methods, Supplementary Results for "Gut microbiome signatures associated with self-reported allergic symptoms among Finnish adults"

### Supplementary information

#### Supplementary Methods

##### Detailed information on the FINRISK 2002 cohort microbiome study

The FINRISK population surveys have been performed every 5 years since 1972. The survey design and methods have been published in detail elsewhere (Salosensaari *et al.* 2020). The FINRISK 2002 study was based on a stratified random sample of the population aged 25-74 years from several geographical areas of Finland. The sampling was stratified by sex, region, and 10-year age groups, each with 250 participants. In North Karelia, Lapland, and the cities of Helsinki and Vantaa, the strata of 65-74-year-old men and women were also sampled, each with 250 participants (Borodulin *et al.* 2018). The original population sample invited to participate in the study was 13,500, with a participation rate of 65.5% (n = 8,798). The survey included a health-related questionnaire, physical examination, collection of stool samples and subsequent metagenome sequencing (Salosensaari *et al.* 2020). The questions on disease history included the following in relation to allergic symptoms: “Have you ever had hay fever or other allergic nasal symptoms?” and “Have you ever had allergic eye symptoms?” (Finn. Inst. Health Welf. THL Finl. 2023). The response choices were “no", “yes, within the past 12 months”, or “yes, over a year ago”. Individuals responding either of “yes” answers to the nasal symptom question were classified into the allergic rhinitis (AR) group, while those responding either of “yes” answers to the eye symptom question were classified into the allergic eye symptoms (AES) group, representing lifetime prevalence of the respective symptoms. We also included a combined group of individuals with either or both symptoms as a Combined Allergies (CA) group. The control group comprised individuals who answered "no" in both symptoms indicating no reported allergic symptoms.

The study protocol of FINRISK 2002 was approved by the Coordinating Ethical Committee of the Helsinki and Uusimaa Hospital District (Ref. 558/E3/2001). All participants signed an informed consent form. The study was conducted in accordance with the World Medical Association’s Declaration of Helsinki on ethical principles. All willing participants in FINRISK 2002 were given a stool sampling kit with detailed instructions. The participants mailed their samples overnight between Monday and Thursday under Finnish winter conditions to the Finnish Institute for Health and Welfare laboratory, where they were stored at -20 °C. The stool samples remained continuously frozen during storage and transport and were transferred in 2017 to the University of California, San Diego, for shotgun metagenomic sequencing (Salosensaari *et al.* 2020). We successfully performed stool shotgun sequencing on n = 7,231 individuals, of whom n = 7,219 had corresponding Greengenes2 profiles. We excluded individuals who did not have linked EHRs of clinical endpoints or who withdrew their consent from the THL Biobank at the time of the study (n = 171), had low read counts (< 50,000, n = 204), who were pregnant at baseline (n = 40), and individuals who had a prescription of antibiotics use defined as ATC code of J01 up to 6 months before baseline (n = 1,241). This resulted in a total n = 5,595. Among those, 5,499 answered the question on allergic rhinitis, and 5,477 answered the question on allergic eye symptoms.

#### Detailed description of taxonomic and functional profiling using shotgun metagenomics

Shotgun metagenomic sequences were trimmed for quality and adapter sequences using Atropos (Didion, Martin, and Collins 2017), and host reads were removed by genome mapping against the human genome assembly GRCh38 with Bowtie2 (Langmead and Salzberg 2012). We mapped filtered reads against two reference databases to support taxonomic and phylogenetic profiling. First, reads were aligned to the Greengenes2 (GG2) reference database (release 2022.10) (McDonald *et al.* 2023), which provides a curated phylogenetic framework specifically tailored for microbiome analysis. Additionally, to support accuracy in the placement of reads within the GG2 phylogeny, we mapped the reads to the Web of Life (WoL) database v2 using Bowtie2. The WoL2 database serves as the comprehensive genomic backbone used in the construction of the GG2 phylogeny. Thus, mapping to WoL2 enables reliable placement of sequences in the broader genomic context that underlies the GG2 tree, while GG2 supports taxonomic assignments and comparative microbiome analysis. The mapped reads were processed by the Woltka pipeline (Q *et al.* 2022). Subsequently, we included the features that overlap with GG2 and assigned the taxonomic information using the QIIME2 (Bolyen *et al.* 2019) q2-greengenes2 plugin. We observed 6,535 taxa as annotated by the pipeline (2 Domains, 90 Phyla, 193 Classes, 533 Orders, 1,015 Families, 3,353 Genera, and 5,965 Species). The taxonomic labels may contain polyphyletic annotation on the taxonomic name where capital letters originate from the relevant label and numbers from the GG2 labels. Certain taxa might have either one or both polyphyletic notations. Although recent taxonomic revisions propose new naming, we retain the names directly provided by the preprocessing pipeline. The functional profiling was performed using HUMAnN3 (Beghini *et al.*, n.d.), yielding 6,535 Uniref90 hits that were further summarized into 298 MetaCyc pathways, and 4,499 KEGG Orthologs (KO). In differential abundance analysis, we only considered taxonomic groups detected with ≥ 1% prevalence at the ≥ 0.1% detection limit. This included 12 Phyla, 16 Classes, 34 Orders, 63 Families, 243 Genera, and 377 Species. Similarly, we analyzed predicted functions detected with ≥ 10% prevalence, retaining 166 MetaCyc pathways and 1,171 KOs.

#### Statistical Analysis

The downstream statistical analysis was performed in R (version 4.3.1) using the TreeSummarizedExperiment (Huang *et al.* 2021) data container with key R/Bioconductor packages, including mia (Borman *et al.* 2025) and miaViz. Observed richness and Shannon diversity were calculated based on species-level counts per sample data. We also included Faith index analysis (Faith 1992), where the diversities are weighted based on phylogeny, as well on species-level counts. We applied rarefaction to account for uneven sequencing depth by randomly subsampling the data 100 times (Schloss 2024) with the mia::addAlpha *niter* parameter. The statistical differences between control and both allergy groups were tested using Wilcoxon rank-sum test, with p-value < 0.05 considered significant. Principal coordinates analyses (PCoA) were based on UniFrac dissimilarity aiming to visualize population variation by the allergy status as it takes into account phylogenetic distances. The UniFrac dissimilarity was calculated using species-level taxa counts. The same rarefaction approach as in the alpha diversity analysis was applied (Schloss 2024). We used Permutational Multivariate Analysis of Variance (PERMANOVA) to quantify differences in microbial community composition between the study groups.

Differential abundance analysis of microbial taxa was conducted using linear models with the MaAsLin2 R package to identify the species that were differentially abundant between groups (Lin, Eggesbø, and Peddada 2022). The differential abundance analysis of the predicted functions was conducted as well with the MaAsLin2. We qualitatively analysed and visualized the enrichment patterns of the pathways using a visualisation and analysis tool Enteropathway (Shiroma *et al.* 2024). The analyses were adjusted for possible confounding factors such as age, BMI, sex, smoking status, and the geographical area in Finland (East and West). The p-values were adjusted using the Holm-Bonferroni method to control for multiple testing, and an adjusted p-value < 0.05 was considered significant.

To evaluate the potential impact of regional variation, additional analyses were performed stratified by region to account for genetic and lifestyle variability (Kerminen *et al.* 2017). We replicated the differential abundance analyses on taxonomic and functional profiles in two sub-populations, Eastern

Finland (n = 3,912), and Western Finland (n = 1,683). The Eastern subset covers the regions of North Karelia, Northern Savonia, Oulu, and Lapland, and the Western subset covers the Turku/Loimaa and Helsinki/Vantaa regions.

### Supplementary Results

#### Volcano plot in the functional predictions analysis

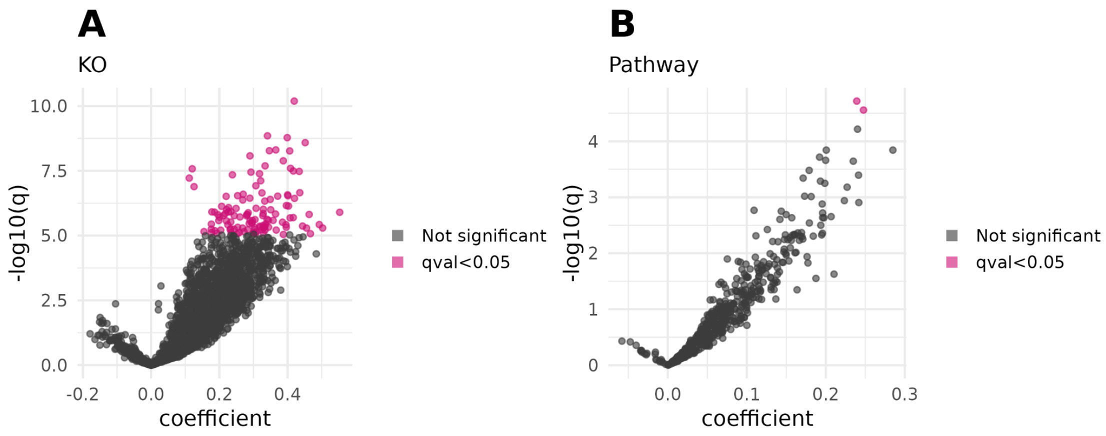


**Figure S1. A** Statistical significance against magnitude of chance in KO functional data, representing several significant KOs enriched in allergic individuals while no significant KOs enriched in controls. The unbalanced coefficient values are shown, arising potentially from coordinated shifts in microbial metabolic potential associated with allergic conditions, or from technical and statistical properties of metagenomic functional profiling. **B** Statistical significance against magnitude of chance in Metacyc pathways.

#### Regional analysis

AR was reported in 33.6% (n = 1,316) and 37.7% (n = 634) of the study population in eastern and western Finland, respectively. For AES, 25.97% (n = 1,016) of the study population in eastern Finland and 31.73% (n = 538) in western Finland reported symptoms. Although the overall trends in taxonomic associations were similar between the whole study population and in the two regional subsets, we did not identify taxa that were consistently significantly associated with allergies across all regions (**Figure S2A**, **Table S4**, **Table S5**).

Several members of the *Clostridia* class (*Ventrisoma faecale*, *Butyricicoccus A 77030 sp900604335*, *Sellimonas intestinalis, Anaerotignum lactatifermentans*), *Fimiplasma intestinipullorum*, and *Adlercreutzia equolifaciens* were significantly enriched in the allergy groups in the overall dataset and in the eastern Finland subset. While these species also showed a similar enrichment trend in the AR group in western Finland, the association was not statistically significant (q > 0.05). Conversely, species such as *Ellagibacter isourolithinifaciens, Desulfovibrio R 446353 piger A, Collinsella* *bouchesdurhonensis* and *Phascolarctobacterium A succinatuens* were significantly less abundant in allergic groups in the Eastern Finland and overall dataset. In the Western Finland and overall dataset, instead, *Flavonifractor plautii, Enterocloster citroniae* were enriched in the allergic groups, whereas *SFTJ01 sp004563195* (*Muribaculaceae* family) and *Prevotella sp003447235* were significantly depleted compared to control. Moreover, *Prevotella copri* and *Prevotella sp002251435* were significantly depleted in the overall dataset compared with controls. Region-stratified analyses showed consistent depletion in AR and CA in western Finland and in AES in eastern Finland. Furthermore, we also identified several species that reached statistical significance in the differential abundance analyses only in either eastern or western Finland, although the qualitative patterns of association were similar across regions. Consistent with previous studies in the same cohort (Ruuskanen *et al.* 2022), these findings suggest that microbial association patterns are largely reproducible across regions, although statistical significance varies.

While functional features showed fewer regionally consistent statistically significant associations, the overall direction of enrichment and depletion remained consistent across the full dataset and both regional subsets (**Figure S2B**, **Table S6**, **Table S7**). Only K06871, an uncharacterized protein, was enriched in all allergic symptom groups in the whole dataset and in Western Finland, as well as in the AES group in Eastern Finland. Other consistent enrichment patterns were detected for K03088, of rpoE a RNA polymerase sigma-70 factor in ECF subfamily, and K07443, methylated DNA protein cysteine transferase related protein, which were both enriched in the whole dataset and in Western Finland. Similarly, K03169 (DNA topoisomerase III), which was significantly enriched in both allergic symptom groups in the whole dataset, was also significantly enriched in allergic groups of Eastern Finland subset. Furthermore, in the pathway analysis for the regional subsets we did not detect statistically significant pathways (**Table S8**, **Table S9**),although the same directional enrichment trends were observed across subsets. In line with the species association trend, these findings suggest that while the functional enrichment trends are broadly consistent across regions, differences in statistical significance may be influenced by variations in sample size and statistical power.

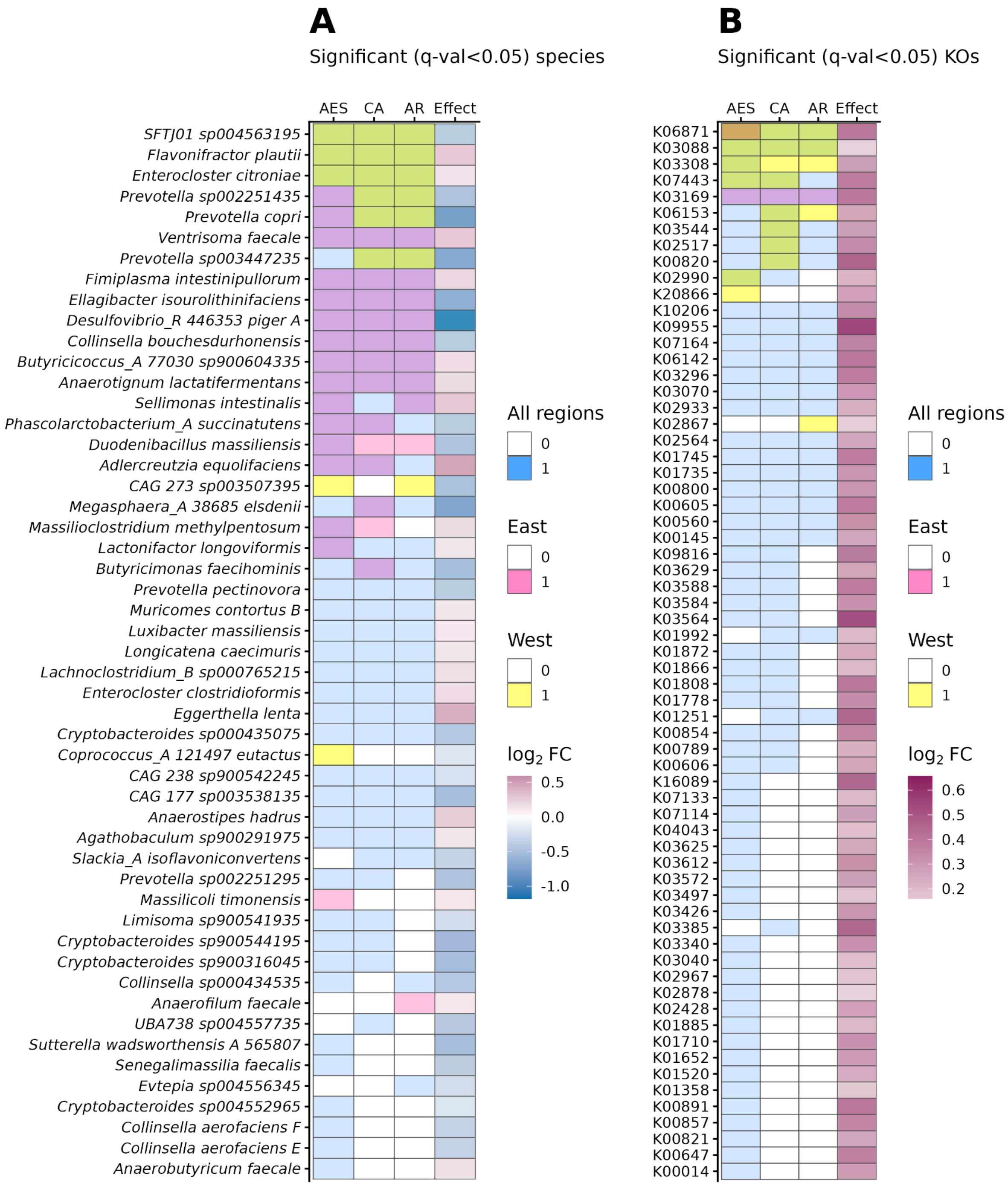


**Figure S2. A** Significant species across all regional sets, where pink represents Eastern subset, blue all samples, and yellow Western subset, while mixture colors violet and green represent significant association in two regions (violet: East and all regions; green West and all regions). The $\log_2$FC represents the $\log_2$ fold change between CA and control group in the whole data, indicating the overall enrichment pattern of the species. **B** Significant KOs across all regional sets. Here brown color in K06871 represents significant association with AES in all three regions. The $\log_2$FC represents the $\log_2$ fold change between CA and control group in the whole data.

#### Differential Progression of Asthma and COPD in Individuals With and Without Allergic Conditions

We found that enriched species associated with allergic rhinitis (AR) and allergic eye symptoms (AES) were also reported to be associated with incident asthma and COPD (Liu *et al.* 2023). Thus, we were interested to assess the proportion of individuals with and without allergic symptoms in developing both chronic lung diseases during the follow-up period available on FINRISK 2002 cohort. We observed a higher proportion of individuals having the allergic symptoms of developing asthma during the follow-up time compared to control individuals. However, the proportions of those developing COPD remain similar between individuals with and without allergic symptoms.

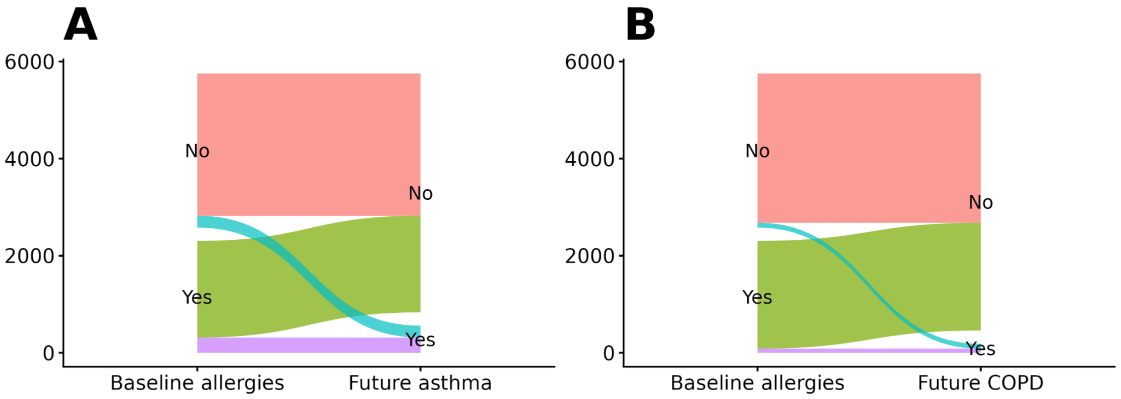


**Figure S3. Asthma and COPD Progression in Allergic vs. Non-Allergic Groups.** Sankey plot illustrating the transition of individuals from baseline allergy status (AR or AES and control) to incident respiratory outcomes (asthma and COPD) over the follow-up period. Each flow represents the proportion of individuals who developed respiratory conditions during follow-up, stratified by baseline self-reported allergy symptoms. **A** Around 14% of those who have baseline allergies develop future asthma, compared to 8% in the control group. **B** Around 3.7% of those who have baseline allergies develop future COPD, compared to 3.1% in the control group.

#### References

Beghini F, McIver LJ, Blanco-Míguez A *et al.* Integrating taxonomic, functional, and strain-level profiling of diverse microbial communities with bioBakery 3. *eLife* n.d.;**10**:e65088. https://doi.org/10.7554/eLife.65088.

Bolyen E, Rideout JR, Dillon MR *et al.* Reproducible, interactive, scalable and extensible microbiome data science using QIIME 2. *Nat Biotechnol* 2019;**37**(8):852–7. https://doi.org/10.1038/s41587-019-0209-9.

Borman T, Benedetti G, Muluh G *et al.* Orchestrating Microbiome Analysis with Bioconductor. Preprint, bioRxiv, 30 Oct. 2025, 2025.10.29.685036. https://doi.org/10.1101/2025.10.29.685036.

Borodulin K, Tolonen H, Jousilahti P *et al.* Cohort Profile: The National FINRISK Study. *Int J Epidemiol* 2018;**47**(3):696–696i. https://doi.org/10.1093/ije/dyx239.

Didion JP, Martin M, Collins FS. Atropos: specific, sensitive, and speedy trimming of sequencing reads. *PeerJ* 2017;**5**:e3720. https://doi.org/10.7717/peerj.3720.

Faith DP. Conservation evaluation and phylogenetic diversity. *Biol Conserv* 1992;**61**(1):1–10. https://doi.org/10.1016/0006-3207(92)91201-3.

Finnish Institute for Health and Welfare (THL), Finland. The National FINRISK Study 1992-2012 - THL. 12 Dec. 2023. https://thl.fi/en/research-and-development/thl-biobank/for-researchers/sample-collections/the-national-finrisk-study-1992-2012 (29 Nov. 2024, date last accessed).

Huang R, Soneson C, Ernst FGM *et al.* TreeSummarizedExperiment: a S4 class for data with hierarchical structure, 9:1246. Preprint, F1000Research, 2 Mar. 2021. https://doi.org/10.12688/f1000research.26669.2.

Kerminen S, Havulinna AS, Hellenthal G *et al.* Fine-Scale Genetic Structure in Finland. *G3 GenesGenomesGenetics* 2017;**7**(10):3459–68. https://doi.org/10.1534/g3.117.300217.

Langmead B, Salzberg SL. Fast gapped-read alignment with Bowtie 2. *Nat Methods* 2012;**9**(4):art. 4. https://doi.org/10.1038/nmeth.1923.

Lin H, Eggesbø M, Peddada SD. Linear and nonlinear correlation estimators unveil undescribed taxa interactions in microbiome data. *Nat Commun* 2022;**13**(1):4946. https://doi.org/10.1038/s41467-022-32243-x.

Liu Y, Teo SM, Méric G *et al.* The gut microbiome is a significant risk factor for future chronic lung disease. *J Allergy Clin Immunol* 2023;**151**(4):943–52. https://doi.org/10.1016/j.jaci.2022.12.810.

McDonald D, Jiang Y, Balaban M *et al.* Greengenes2 unifies microbial data in a single reference tree. *Nat Biotechnol* 27 Jul. 2023:1–4. https://doi.org/10.1038/s41587-023-01845-1.

Q Z, S H, A G *et al.* Phylogeny-Aware Analysis of Metagenome Community Ecology Based on Matched Reference Genomes while Bypassing Taxonomy. *mSystems* 2022;**7**(2). https://doi.org/10.1128/msystems.00167-22.

Ruuskanen MO, Erawijantari PP, Havulinna AS *et al.* Gut Microbiome Composition Is Predictive of Incident Type 2 Diabetes in a Population Cohort of 5,572 Finnish Adults. *Diabetes Care* 2022;**45**(4):811–8. https://doi.org/10.2337/dc21-2358.

Salosensaari A, Laitinen V, Havulinna AS *et al.* Taxonomic Signatures of Long-Term Mortality Risk in Human Gut Microbiota. *medRxiv* 13 Jan. 2020:2019.12.30.19015842. https://doi.org/10.1101/2019.12.30.19015842.

Schloss PD. Rarefaction is currently the best approach to control for uneven sequencing effort in amplicon sequence analyses. *mSphere* 2024;**9**(2):e00354-23. https://doi.org/10.1128/msphere.00354-23.

Shiroma H, Darzi Y, Terajima E *et al.* Enteropathway: the metabolic pathway database for the human gut microbiota. *Brief Bioinform* 2024;**25**(5):bbae419. https://doi.org/10.1093/bib/bbae419.
